# Exploring Burnout and Mindfulness among Medical Researchers: A Global Cross-Sectional Survey

**DOI:** 10.64898/2026.02.24.26346825

**Authors:** Jeremy Y. Ng, Niveen Syed, Gabriela Melendez, Mirela I. Bilc, Anna Katharina Koch, Holger Cramer

## Abstract

Burnout, a state of chronic exhaustion often characterized by feelings of emotional exhaustion, cognitive and emotional dysregulation, and psychological distancing, is an increasingly recognized issue within most professions. This syndrome results in diminished job satisfaction, strained interpersonal relationships, and decreased well-being. Socio-demographic factors have been shown to play a role in burnout risk, while trait mindfulness has been identified as an effective method to mitigate it. This study aimed to identify the prevalence of burnout risk and its relationship with mindfulness and socio-demographics among medical researchers. An anonymous, online, cross-sectional survey was administered to corresponding authors published in MEDLINE. The survey consisted of screening and socio-demographic questions, as well as validated assessment tools (i.e., shortened work-related Burnout Assessment Tool [BAT-12] and shortened Freiburg Mindfulness Inventory [FMI-14]). Responses were analysed according to the BAT and FMI guidelines, alongside regression analyses. A total of 1,732 participants completed the survey, yielding a response rate of 1.88%. Overall, 38.8% of participants were at risk or at very high risk of burnout, and the mean mindfulness score was 37.51. Multiple linear regression analysis indicated that sex, age, and employment status were significant predictors of burnout risk, while age and region significantly predicted mindfulness. Hierarchical regression analysis showed that, after controlling for socio-demographic variables, mindfulness was a strong and independent negative predictor of burnout risk. These findings on burnout risk and the influence of mindfulness and socio-demographics could guide future research in developing tailored interventions and policies that improve the well-being of medical researchers.

## Background

Mental health refers to a state of well-being in which an individual can productively contribute to their community and effectively cope with stress. [1] Burnout, a psychological state caused by prolonged exposure to interpersonal stressors, typically related to work (e.g., job insecurity, increased workload), can diminish mental health. [2,3] It is characterized by three primary symptoms: emotional exhaustion, cognitive and emotional dysregulation, and psychological distancing. [3] As these symptoms worsen, they can exacerbate physical and other mental health problems, resulting in a vicious cycle of declining well-being. Recent research has identified a reciprocal relationship between burnout and clinical depression, along with anxiety disorders developing or worsening due to prolonged stress and burnout. [4–7] Collectively, these effects can adversely impact well-being and impair cognitive functioning, leading to decreased productivity, strained relationships, and reduced quality of life. [3,8–10]

Notably, the risk of burnout is not evenly distributed across populations. Growing evidence indicates that demographic characteristics, including age, gender, ethnicity, career type, career stage, and geographic location, significantly shape vulnerability to burnout. [11–15] Structural and institutional factors often interact with these characteristics to amplify stress and limit access to protective resources. [12,16,17] For example, women consistently report higher levels of burnout than men across professional settings. These disparities are thought to arise from greater exposure to bias and microaggressions, disproportionate role expectations and responsibilities, fewer leadership and advancement opportunities, and limited access to specialized support systems, resulting in chronic stress and elevated burnout risk. [11,12,16]

Within this broader context, researchers are particularly susceptible to burnout due to the demanding nature of their profession. Long working hours, job insecurity due to grant-funding instability, and inadequate institutional support within academic and research settings have consistently been associated with higher burnout, particularly among early- and mid-career researchers. [2,18–23] Even before the COVID-19 pandemic, an estimated 30% to 40% of researchers reported experiencing mental health problems, already exceeding the risk rate observed in the general population. Since then, this risk has significantly increased as working conditions have deteriorated. [2,24] Ultimately, the competitive and unstable nature of academic work continues to exacerbate burnout among researchers, contributing to their anxiety and depression. [2,23,25]

Medical researchers face additional pressures within this environment. The demands of a pervasive ‘publish or perish’ culture place medical researchers under significant pressure to produce rapid results while navigating evolving health priorities, work insecurity, competitive funding, and strict deadlines. [20,21,26–31] A nationwide study in the Netherlands found that 24% of medical professors exhibited symptoms of burnout solely due to publication pressure. [27] This urgency to publish can also contribute to scientific misconduct and inaccuracies, further burdening medical researchers as they work to combat the spread of misinformation. [20,28,32] These challenges contribute to heightened distress and burnout among medical researchers, leading to reduced well-being. [26,27,30]

Clinician-researchers face the added challenge of balancing clinical responsibilities with research activities, providing patient care alongside budgeting, grant-writing, and personnel management. The combined demands of treating patients while producing relevant studies create a unique set of pressures that can significantly impact well-being. [21,22,26–28,33,34] Even when physician-researchers received considerable support from the National Institutes of Health, work-related burnout was reported in 41.4% of women and 31.5% of men. [33] This demanding schedule leads to physical and mental exhaustion and puts immense strain on interpersonal relationships, contributing to burnout and declining well-being within this group.

While burnout affects medical researchers individually, it also has large-scale consequences. In academic medicine and clinical research, burnout increases intentions to quit, creating significant barriers to effective knowledge dissemination and patient care. These barriers include lower-quality and reduced research output, as well as hindered sharing of important scientific discoveries. [2,22,29–31] Additionally, the emotional exhaustion associated with burnout can ultimately compromise the quality of care provided to patients. [2,22,33,34] Addressing these challenges is essential to preserving the integrity and effectiveness of medical research and clinical practice and supporting the well-being of those who conduct it.

Fortunately, adaptive coping mechanisms, defined as cognitive and behavioural processes that manage the demands placed on an individual, can help mitigate the effects of burnout and improve mental health. [35] Among the strategies for coping with burnout, mindfulness, a trait characterized by self-understanding and present-moment awareness, stands out as being particularly effective. Research indicates that mindfulness helps with self-awareness and emotional regulation, reducing rumination and negative thought patterns that can exacerbate stress. [35] Additionally, individuals with higher dispositional mindfulness report an improved ability to identify and manage work-related stress. They also experience reduced emotional exhaustion, which is a key component of burnout. [36–39] Ultimately, mindfulness can enhance psychological resilience, mood, and self-efficacy, enabling individuals to navigate workplace demands and maintain well-being even in high-pressure environments. [35,39–41]

While burnout has been extensively examined among physicians, nurses, and other healthcare professionals engaged in direct patient care [11,15,34,42–47], burnout among medical researchers remains relatively understudied, particularly regarding how socio-demographic factors and trait mindfulness may intersect to influence burnout risk. Existing studies often examine these factors in isolation (e.g., focusing on early- or mid-career researchers separately rather than comparatively [18,19]), considering only one socio-demographic factor at a time (e.g., biomedical academic researchers and faculty [30]), or conducting nationwide rather than international studies [20,27,28]. As a result, the risk of burnout among medical researchers is not well characterized, and gaps remain in understanding how specific socio-demographic characteristics and trait mindfulness interact to shape burnout risk.

Leveraging a cross-sectional survey, our objective was to identify burnout risk and trait mindfulness among medical researchers using validated assessment tools. We also aimed to examine the relationships between burnout, mindfulness, and socio-demographic factors, specifically whether socio-demographic characteristics predicted burnout risk and mindfulness and whether mindfulness uniquely contributed to the prediction of burnout risk after controlling for demographic factors. Insights from this study can guide future research aimed at identifying and developing tailored interventions to improve the well-being of medical researchers worldwide.

## Methods

### Transparency Statement

Before commencing this project, clearance and approval were obtained from the University Hospital Tübingen Research Ethics Board (REB #: 217/2025BO2). The study protocol was registered and made available on the Open Science Framework (OSF) [48] before participant recruitment, while study materials and data were anonymized and shared via OSF once they were available [49]. No personal data was collected or processed in this study, and IP addresses were not stored. Consent for data collection, storage, use, and sharing was obtained through a consent form. Participants could withdraw from the online survey at any time, and incomplete surveys were not analysed. The manuscript was reported in accordance with STrengthening the Reporting of OBservational studies in Epidemiology (STROBE) [50]. The resulting manuscript was posted on a preprint server before submission to a journal.

### Study Design

An anonymous, online, cross-sectional survey was administered to corresponding authors identified as having published in MEDLINE-indexed journals between December 13, 2023, and January 25, 2024 (to ensure a high response rate from active researchers). A systematic approach was used to identify participants, beginning with the randomization of a predefined list of NLM journal IDs for journal selection, followed by an automated search strategy to identify researchers from articles published in the listed journals. This approach yielded approximately 100,000 unique email addresses. All identified researchers were invited to participate, and a screening question confirmed eligibility to complete the survey.

### Sampling Framework

A comprehensive list of MEDLINE-indexed journals and their NLM IDs was obtained from inputting “currentlyindexed [All Fields] AND currentlyindexedelectronic [All Fields]” into the search bar of the NLM Catalogue, totalling 5043 journals as of January 2024. A search strategy in OVID MEDLINE was developed to identify NLM IDs and limit the study’s focus to journals published between December 13, 2023, and January 25, 2024 (see **Appendix 1**). Authors of all medical article types were included. Journals only published in English were sampled. To obtain author names, affiliated institutions, and email addresses, the PMID numbers linked to these articles were exported from OVID as a .csv file and processed through an R script that was developed using easyPubMed. [51] The results were compiled into a master list and reviewed for errors or duplicates before participant recruitment and survey administration.

### Participant Recruitment

Only researchers identified through the sampling framework as published medical researchers were invited to complete the closed survey. Eligible participants were those whose research contributed to the field of medicine and who had either completed a terminal degree in their field (e.g., PhD, MD, or equivalent) or had more than 5 years of experience in a research-focused role (e.g., research coordinator).

The survey was created and distributed via SurveyMonkey software. [52] The emails containing the invitation to complete the anonymous online survey also included an authorized recruitment script, an explanation of the study and its goals, and an informed consent form (see https://osf.io/eawd8/files/qevwf). The data underwent a cleaning process to correct names that were improperly extracted, remove email addresses that were duplicated, and eliminate names with invalid email addresses. The survey was open from September 22, 2025, to October 27, 2025. Participants received reminder emails during the first, second, and third weeks following the original invitation email. The survey was closed two weeks following the final reminder, giving respondents a total of five weeks to complete the survey. The initial recruitment and follow-up reminder emails can be found at https://osf.io/eawd8/files/6kysw. Participation was not mandatory, and financial compensation was not provided in this study.

### Survey Design

The survey first asked participants a screening question to confirm participant eligibility as medical researchers, followed by general socio-demographic questions. Participants were then asked to complete two validated assessment tools: the work-related 12-item Burnout Assessment Tool (BAT-12) [53] and the 14-item Freiburg Mindfulness Instrument (FMI-14) [54] to measure burnout risk and trait mindfulness, respectively.

The BAT-12 is the shortened version of the original 23-item Burnout Assessment Tool (BAT), which was developed through extensive practitioner interviews and a review of existing burnout scales. [3] Psychometric studies across multiple countries have shown that the BAT has high reliability, internal consistency, and convergent validity with other burnout instruments, while remaining distinct from related constructs such as work engagement or job boredom. [3,55,56] It also uses clinically validated cut-off scores to identify individuals with severe burnout, with thresholds derived from samples of individuals who, according to expert assessments by psychologists, general practitioners, and occupational physicians, are clinically diagnosed with burnout. [3] The BAT-12 retains these key features while reducing survey completion time, an important consideration for busy medical researchers. [53,55–58] Additionally, the work-related version of BAT-12 specifically assesses burnout risk in the professional context, aligning directly with our study’s focus on occupational stressors. [53]

The FMI-14 is the short form of the original 30-item Freiburg Mindfulness Inventory (FMI), which was developed through expert interviews and literature review, and tested with participants attending meditation retreats. [54] The FMI has been validated across diverse populations, from individuals without meditation experience to clinical patients, and demonstrates high internal consistency. The FMI-14 maintains these psychometric strengths and broad applicability while decreasing survey completion time, making it a practical choice for our study. [54]

The screening and demographic questions were presented in a multiple-choice format, while the assessment tools for burnout risk and trait mindfulness were presented in their original format, a Likert scale, to ensure scale validity. We anticipated that the survey would take less than 10 minutes to complete. A copy of the survey is provided in **Appendix 2** and can be found at https://osf.io/eawd8/files/zeufr.

### Data Analysis

Data from participants who completed the BAT-12 and FMI-14 in full were analysed. Participant demographics were summarized using frequencies and percentages.

Participant responses for burnout and mindfulness were first analysed individually to ensure accurate categorization, then aggregated to determine overall frequencies, percentages, and other basic statistics. Burnout scoring followed BAT guidelines: total scores were calculated by summing response-specific values and dividing by the number of items, with the same procedure applied to each dimension. Scores were then categorized as no risk, at risk, or very high risk based on established thresholds. [3,53] Mindfulness was measured using the FMI protocol, with total scores obtained by adding item-specific values. [54]

Internal consistency of the BAT and FMI was assessed using Cronbach’s alpha. Multiple linear regression analyses were used to assess the predictive role of socio-demographic characteristics (e.g., age, gender, career stage, research area) in relation to both burnout risk and trait mindfulness. Burnout risk and mindfulness were each analysed as a continuous dependent variable. Socio-demographic factors were defined as categorical independent variables, which were dummy coded with the most frequent response category as the reference.

A Pearson correlation was conducted to examine the association between burnout (BAT total score) and mindfulness (FMI total score). To further examine this relationship, a hierarchical linear regression analysis was conducted with burnout risk as the outcome variable. Demographic variables were entered in the first step, followed by mindfulness in the second step, allowing for the assessment of the unique predictive contribution of mindfulness after controlling for demographic factors.

Model fit was assessed using R², which measured how well the independent variables explained burnout and trait mindfulness. Statistical significance was assessed using a threshold of p<0.05, and all analyses were conducted using R (version 4.5.0) [59] and RStudio (version 2024.12.1.563) [60] (see **Appendix 3**). A power analysis was not conducted, as it was not required for our purposive sampling approach, and our extensive dataset of approximately 100,000 unique emails ensured a large sample size.

## Results

### Socio-demographics

A total of 99636 survey invitations were emailed to identified corresponding authors, of which 7727 bounced back. Of the 91909 invitations that were successfully delivered, 1732 completed the survey in full (see https://osf.io/eawd8/files/a7rcp), yielding a response rate of 1.88%. When only considering the 49386 invitations that were opened, a response rate of 3.50% was achieved. The survey took approximately nine minutes to complete.

Respondents represented a range of career stages, with the majority being senior researchers with more than ten years of experience since starting their career post-PhD, MD, or an equivalent (n=1041, 60.1%). Mid-career researchers with five to ten years of experience included 387 participants (22.3%), while early-career researchers with less than five years of experience comprised 294 participants (17.0%). Participants who identified as male or female were similarly represented, comprising 50.2% (n=869) and 48.7% (n=843), respectively. Most respondents were between the ages of 36 and 55 (n=1,069, 61.7%), with 24.4% over 55 (n=422) and 13.9% under 36 (n=241).

The majority of respondents were employed in Europe (n=621, 35.9%) or the Americas (n=601, 34.7%), with smaller proportions working in South-East Asia (n=186, 10.7%), the Western Pacific (n=132, 7.6%), Africa (n=78, 4.5%), and the Eastern Mediterranean (n=69, 4.0%). Most respondents reported that English was not their primary language (n=948, 54.73%), compared with 777 respondents (44.86%) who indicated that it was.

Regarding professional roles, nearly half were faculty members at universities or academic institutions (n=1159, 48.4%), followed by clinician researchers (n=402, 16.8%), academic research staff (n=354, 14.8%), and journal editors or editorial board members (n=174, 7.3%). Less than three percent of respondents selected roles as consultants, government researchers, or industry professionals (e.g., pharmacy, scholarly communication, and third-sector organizations). The most frequently selected research areas were clinical research (n=966, 31.49%), followed by epidemiological research (n=391, 12.74%), preclinical in vitro research (n=374, 12.19%), and preclinical in vivo research (n=354, 11.54%). Other research areas included health services research (n=327, 10.66%), methods research (n=281, 9.16%), and health systems research (n=233, 7.59%).

A wide range of research disciplines was reported. The most represented area was public health (n=458, 12.98%), followed by biochemistry, cell biology, and genetics (n=290, 8.22%), the nervous system (n=209, 5.92%), microbiology and immunology (n=197, 5.58%), and the cardiovascular system (n=161, 4.56%). Other disciplines included general medicine/health professions (n=155, 4.39%), pediatrics (n=142, 4.02%), physiology (n=140, 3.97%), pharmacology (n=139, 3.94%), and psychiatry (n=139, 3.94%). Smaller proportions of respondents (3.85% and lower) reported specialties across organ systems (e.g., musculoskeletal, respiratory, digestive, endocrine), clinical subspecialties (e.g., surgery, radiology, obstetrics, dermatology), and applied health sciences (e.g., nursing, dentistry, health facilities management). Most respondents indicated full-time employment (n=1526, 88.11%), while smaller proportions reported working part-time (n=123, 7.10%) or being self-employed (n=28, 1.62%). A summary of anonymized, aggregated participant socio-demographics is presented in **Table 1**, while a complete list is available at https://osf.io/eawd8/files/vkz42.

**Table 1:** Participant Demographics.

| Demographics | Participant Characteristics | Responses (n (%)) |
| --- | --- | --- |
| Career Stage | Early career researcher (<5 years of formally starting your career post PhD, MD, or an equivalent) | 294 (16.97%) |
|  | Mid-career researcher (5-10 years of starting your career post PhD, MD, or an equivalent) | 387 (22.34%) |
|  | Senior researcher (>10 years of starting your career post PhD, MD, or an equivalent) | 1041 (60.10%) |
|  | Prefer not to disclose | 10 (0.58%) |
|  |  | **N=1732** |
| Position | Faculty member at a university/academic institution | 1159 (48.43%) |
|  | Academic research staff (e.g., research coordinator at a university/academic institution) | 354 (14.79%) |
|  | Government researcher | 67 (2.80%) |
|  | Clinician researcher | 402 (16.80%) |
|  | Pharmaceutical industry/company researcher or employee | 24 (1.00%) |
|  | Scholarly communications employee (e.g., scholarly journals/publishing, medical writing) | 14 (0.59%) |
|  | Journal editor or editorial board member | 174 (7.27%) |
|  | Consultant (e.g., own or employed by research consulting firm) | 71 (2.97%) |
|  | Third sector (e.g., NGO, non-profit) | 31 (1.30%) |
|  | Prefer not to disclose | 10 (0.42%) |
|  | Other | 87 (3.64%) |
|  |  | **N=2393<sup>a</sup>** |
| Sex | Female | 843 (48.67%) |
|  | Male | 869 (50.17%) |
|  | Prefer not to disclose | 17 (0.98%) |
|  | Prefer to self-describe | 3 (0.17%) |
|  |  | **N=1732** |
| Age | 18-24 | 7 (0.40%) |
|  | 25-35 | 234 (13.51%) |
|  | 36-45 | 561 (32.39%) |
|  | 46-55 | 508 (29.33%) |
|  | 56-65 | 289 (16.69%) |
|  | 66-70 | 69 (3.98%) |
|  | 71-75 | 34 (1.96%) |
|  | 76-80 | 16 (0.92%) |
|  | 80+ | 6 (0.35%) |
|  | Prefer not to disclose | 8 (0.46%) |
|  |  | **N=1732** |
| Country of Employment | Africa | 78 (4.50%) |
|  | Americas | 601 (34.70%) |
|  | Eastern Mediterranean | 69 (3.98%) |
|  | Europe | 621 (35.85%) |
|  | South-East Asia | 186 (10.74%) |
|  | Western Pacific | 132 (7.62%) |
|  | Prefer not to disclose | 45 (2.60%) |
|  |  | **N= 1732** |
| English as Their Primary Language | Yes | 777 (44.86%) |
|  | No | 948 (54.73%) |
|  | Prefer not to disclose | 7 (0.4%) |
|  |  | **N=1732** |
| Primary Research Area | Clinical research | 966 (31.49%) |
|  | Preclinical research – in vivo | 354 (11.54%) |
|  | Preclinical research – in vitro | 374 (12.19%) |
|  | Health systems research | 233 (7.59%) |
|  | Health services research | 327 (10.66%) |
|  | Methods research | 281 (9.16%) |
|  | Epidemiological research | 391 (12.74%) |
|  | Prefer not to disclose | 10 (0.33%) |
|  | Other | 132 (4.3%) |
|  |  | **N=3068<sup>a</sup>** |
| Research Focus / Discipline | Human Anatomy | 29 (0.82%) |
|  | Physiology | 140 (3.97%) |
|  | Biochemistry. Cell Biology and Genetics | 290 (8.22%) |
|  | Pharmacology | 139 (3.94%) |
|  | Microbiology and Immunology | 197 (5.58%) |
|  | Parasitology. Disease Vectors | 43 (1.22%) |
|  | Clinical Laboratory Pathology | 39 (1.11%) |
|  | Pathology | 69 (1.96%) |
|  | General Medicine. Health Professions | 155 (4.39%) |
|  | Public Health | 458 (12.98%) |
|  | Practice of Medicine | 118 (3.34%) |
|  | Communicable Diseases | 89 (2.52%) |
|  | Medicine in Selected Environments | 26 (0.74%) |
|  | Musculoskeletal System | 136 (3.85%) |
|  | Respiratory System | 86 (2.44%) |
|  | Cardiovascular System | 161 (4.56%) |
|  | Hemic and Lymphatic Systems | 41 (1.16%) |
|  | Digestive System | 96 (2.72%) |
|  | Urogenital System | 53 (1.5%) |
|  | Endocrine System | 75 (2.13%) |
|  | Nervous System | 209 (5.92%) |
|  | Psychiatry | 139 (3.94%) |
|  | Radiology. Diagnostic Imaging | 78 (2.21%) |
|  | Surgery. Wounds and Injuries | 87 (2.47%) |
|  | Gynecology. Andrology | 39 (1.11%) |
|  | Obstetrics | 44 (1.25%) |
|  | Dermatology. Integumentary System | 26 (0.74%) |
|  | Pediatrics | 142 (4.02%) |
|  | Geriatrics | 55 (1.56%) |
|  | Dentistry. Oral Surgery | 43 (1.22%) |
|  | Otolaryngology | 22 (0.62%) |
|  | Ophthalmology | 42 (1.19%) |
|  | Hospitals and Other Health Facilities | 77 (2.18%) |
|  | Nursing | 70 (1.98%) |
|  | History of Medicine. Medical Miscellany | 16 (0.45%) |
|  |  | **N=3529 <sup>a</sup>** |
| Employment Status | Full-time | 1526 (88.11%) |
|  | Part-time | 123 (7.1%) |
|  | Self-employed | 28 (1.62%) |
|  | On leave | 5 (0.29%) |
|  | Unemployed | 8 (0.46%) |
|  | Prefer not to disclose | 6 (0.35%) |
|  | Other | 36 (2.08%) |
|  |  | **N=1732** |

<sup>a</sup> Participants were able to choose more than one option.

### Prevalence of Burnout and Mindfulness

Both the BAT total score and FMI total score demonstrated excellent internal consistency, with Cronbach’s alpha=0.883 and 0.884, respectively.

Overall, nearly 40% of participants were at risk or at very high risk of burnout (n=672), including 21.0% at risk (n=364) and 17.8% at very high risk (n=308). When examining specific dimensions of burnout, mental distancing affected slightly more than half of the participants (n=904, 52.2%), with 37.4% at risk (n=647) and 14.8% at very high risk (n=257). Exhaustion impacted 36.9% of participants (n=639), including 12.2% at risk (n=211) and 24.7% at very high risk (n=428). Emotional impairment affected nearly one-third of participants (n=514, 29.7%), with 19.6% at risk (n=339) and 10.1% at very high risk (n=175). In contrast, cognitive impairment was less prevalent, with 8.3% of participants at risk (n=144) and 7.3% at very high risk (n=126), and the majority (84.4%, n=1462) observing no risk. A summary of participant burnout risks is provided in **Table 2**.

**Table 2:** Participant Burnout Risk (Cut-off)

| **Burnout Type** | **Burnout Risk** | **Responses (Number)** | **Responses (%)** |
| --- | --- | --- | --- |
| **Average Burnout Risk Score** | No risk of burnout | 1060 | 61.20 |
|  | At risk of burnout | 364 | 21.02 |
|  | Very high risk of burnout | 308 | 17.78 |
|  | Total | 1732 | 100.00 |
| **Exhaustion** | No risk of burnout | 1093 | 63.11 |
|  | At risk of burnout | 211 | 12.18 |
|  | Very high risk of burnout | 428 | 24.71 |
|  | Total | 1732 | 100.00 |
| **Mental Distancing** | No risk of burnout | 828 | 47.81 |
|  | At risk of burnout | 647 | 37.36 |
|  | Very high risk of burnout | 257 | 14.84 |
|  | Total | 1732 | 100.00 |
| **Cognitive Impairment** | No risk of burnout | 1462 | 84.41 |
|  | At risk of burnout | 144 | 8.31 |
|  | Very high risk of burnout | 126 | 7.27 |
|  | Total | 1732 | 100.00 |
| **Emotional Impairment** | No risk of burnout | 1218 | 70.32 |
|  | At risk of burnout | 339 | 19.57 |
|  | Very high risk of burnout | 175 | 10.10 |
|  | Total | 1732 | 100.00 |

The trait mindfulness scores of participants ranged from 14 to 56. The sample had a mean score of 37.51 (SD=7.64), a median score of 38, and a mode of 39. A complete list of participant mindfulness scores can be found at https://osf.io/eawd8/files/vkz42.

### Relationships between Burnout, Mindfulness, and Socio-demographic Variables

A multiple linear regression was conducted to examine whether socio-demographic factors, including career stage, sex, age, region, and employment status, predicted burnout scores (see **Table 3**). The overall model was statistically significant, F(26, 1702) 4.83, p<.001, and explained 6.9% of the variance in burnout scores (R²=.069, adjusted R²=.055). Sex, age and employment status emerged as significant predictors, while career stage and region did not. Female participants reported significantly higher burnout compared to male participants (p<.003). Regarding age, older participants aged 66–70, 71–75, 76–80, and 80+ reported significantly lower burnout scores compared to the reference group aged 46-55: 66–70 years (p=.010), 71–75 years (p<.001), 76–80 years (p=.011), and 80+ (p=.004). In terms of employment status, part-time workers reported significantly lower burnout compared to full-time workers (p=.034).

**Table 3:** Multiple Regression Predicting Burnout and Mindfulness.

|  | Burnout |  |  |  |  | Mindfulness |  |  |  |
| --- | --- | --- | --- | --- | --- | --- | --- | --- | --- |
|  | B | SE | t | p |  | B | SE | t | p |
| (Intercept) | 2.318 | 0.037 | 62.62 | **< .001** |  | 37.402 | 0.468 | 79.909 | **< .001** |
| Career stage |  |  |  |  |  |  |  |  |  |
| Early career | −0.001 | 0.055 | −0.024 | .981 |  | −0.959 | 0.696 | −1.379 | .168 |
| Mid-career | 0.007 | 0.043 | 0.167 | .867 |  | −0.918 | 0.541 | −1.696 | .090 |
| Prefer not to disclose | −0.368 | 0.198 | −1.858 | .063 |  | 0.519 | 2.506 | 0.207 | .836 |
| Sex |  |  |  |  |  |  |  |  |  |
| Female | 0.088 | 0.030 | 2.984 | **.003** |  | −0.665 | 0.373 | −1.782 | .075 |
| Prefer not to disclose | 0.317 | 0.172 | 1.845 | .065 |  | −4.222 | 2.170 | −1.946 | .052 |
| Age |  |  |  |  |  |  |  |  |  |
| 18–24 | 0.410 | 0.235 | 1.749 | .081 |  | −8.930 | 2.966 | −3.011 | **.003** |
| 25–35 | 0.069 | 0.062 | 1.116 | .264 |  | −0.787 | 0.786 | −1.002 | .317 |
| 36–45 | 0.071 | 0.041 | 1.748 | .081 |  | −0.784 | 0.516 | −1.520 | .129 |
| 56–65 | −0.082 | 0.044 | −1.861 | .063 |  | 0.607 | 0.560 | 1.083 | .279 |
| 66–70 | −0.201 | 0.079 | −2.559 | **.011** |  | 1.781 | 0.995 | 1.790 | .074 |
| 71–75 | −0.546 | 0.110 | −4.968 | **< .001** |  | 3.388 | 1.389 | 2.440 | **.015** |
| 76–80 | −0.413 | 0.163 | −2.529 | **.012** |  | 2.519 | 2.066 | 1.219 | .223 |
| 80+ | −0.764 | 0.263 | −2.903 | **.004** |  | 5.363 | 3.327 | 1.612 | .107 |
| Prefer not to disclose | −0.192 | 0.246 | −0.778 | .436 |  | 5.568 | 3.112 | 1.789 | .074 |
| Region |  |  |  |  |  |  |  |  |  |
| South-East Asia | 0.053 | 0.050 | 1.057 | .291 |  | 2.365 | 0.636 | 3.719 | **< .001** |
| Americas | 0.057 | 0.034 | 1.665 | .096 |  | 0.946 | 0.436 | 2.169 | **.030** |
| Western Pacific | 0.068 | 0.057 | 1.183 | .237 |  | 0.739 | 0.724 | 1.020 | .308 |
| Eastern Mediterranean | 0.045 | 0.076 | 0.587 | .558 |  | 0.210 | 0.963 | 0.218 | .827 |
| Africa | −0.057 | 0.072 | −0.785 | .432 |  | 2.617 | 0.914 | 2.864 | **.004** |
| Prefer not to disclose | 0.013 | 0.093 | 0.134 | .893 |  | 1.524 | 1.179 | 1.292 | .196 |
| Employment Status |  |  |  |  |  |  |  |  |  |
| Unemployed | 0.000 | 0.214 | 0.001 | .999 |  | −2.556 | 2.709 | −0.944 | .345 |
| Self-employed | −0.082 | 0.116 | −0.712 | .477 |  | 1.369 | 1.462 | 0.936 | .349 |
| Part-time | −0.122 | 0.058 | −2.120 | .034 |  | 0.909 | 0.729 | 1.247 | .213 |
| On leave | −0.053 | 0.286 | −0.184 | .854 |  | 4.007 | 3.620 | 1.107 | .268 |
| Prefer not to disclose | 0.425 | 0.247 | 1.720 | .086 |  | −2.759 | 3.122 | −0.884 | .377 |
| Others | −0.162 | 0.109 | −1.494 | .135 |  | 1.700 | 1.373 | 1.238 | .216 |

Note: Reference categories – Career Stage: Senior researcher; Sex: Male; Age: 46–55; Region: Europe; Employment Status: Full-time; statistically significant effects are bolded

A multiple linear regression was conducted to examine whether socio-demographic factors, including career stage, sex, age, region, and employment status, predicted mindfulness scores (see **Table 3**). The overall model was statistically significant, F(26, 1702)=3.77, p<.001, and explained 5.4% of the variance in mindfulness scores (R²=.054, adjusted R²=.040). Age and region emerged as significant predictors, while career stage, sex and employment status did not. Compared to the 46-55 years reference group, younger participants aged 18-24 years reported significantly lower mindfulness (p=.003), while older participants aged 71-75 years reported higher mindfulness. Participants working in South-East Asia (p<.001), the Americas (p=.030), and Africa (p=.004) reported higher mindfulness scores than those from Europe.

Results of the Pearson correlation revealed a moderate to large, negative relationship between burnout and mindfulness, r=–0.49, p<.001, 95% CI [–0.53, – 0.45]. This indicates that higher levels of burnout were associated with lower levels of mindfulness. Building on this association, a hierarchical linear regression analysis was conducted to examine whether mindfulness predicted burnout risk after controlling for sociodemographic factors. Demographic variables were entered in Step 1 and accounted for 3.4% of the variance in burnout risk (R²=0.034, F(5, 1723)=12.09, p<.001). In Step 2, the inclusion of mindfulness significantly improved the model, explaining an additional 22% of the variance in burnout risk (ΔR²=.222, ΔF(1, 1722)=514.89, p<.001). Mindfulness emerged as a significant negative predictor of burnout risk (B=-0.038, SE=0.002, β=-0,478, t=-22.691, p<.001), indicating that higher levels of mindfulness were associated with lower burnout risk independently of sociodemographic characteristics.

## Discussion

Using a global cross-sectional survey and validated assessment tools, this study examined the risk of burnout and trait mindfulness among medical researchers. It also measured the correlation between these constructs and the moderating influence of socio-demographic characteristics.

This study found that a substantial proportion of medical researchers were at high or very high risk of burnout (n=672, 38.8%), as determined by clinically validated cut-off scores. [3,53] This prevalence is consistent with literature documenting high levels of burnout in academic medicine and research-intensive environments. [2,3,14,18,19,22,27–31,33] Among burnout dimensions, mental distancing was most pronounced, with over half of participants classified as at risk or very high risk. These findings align with other research that indicates disengagement to be a common response to burnout and chronic occupational stress in high-pressure settings. This often arises from unsustainable workloads, career progression uncertainty, and limited institutional support. [2,3,9,16,17,21–23,28,31]

Moreover, emotional impairment was observed in nearly one-third of the sample, while exhaustion affected more than one-quarter of participants. Previous studies have shown that emotional strain and exhaustion are increasingly prevalent in research settings, where prolonged working hours, sustained publication pressures, and limited recovery time are common stressors. [27–31] Interestingly, however, cognitive impairment was comparatively less prevalent, suggesting that many medical researchers retain functional cognitive capacity. Although burnout has been associated with cognitive decline, such impairments may emerge later or be masked by compensatory strategies employed by professionals, such as investing additional time or effort into urgent tasks. [8,9]

Aligning with the broader burnout research [11,12], our results also show that female participants reported higher burnout than males. The observed association between age and burnout, whereby older participants reported significantly lower burnout levels than middle-aged participants, is consistent with a substantial body of prior research suggesting that early career individuals tend to experience higher burnout, potentially due to greater work–family conflict, career uncertainty, and higher performance pressures during these life stages. [61] Part-time workers’ lower burnout scores may reflect reduced job demands or more flexibility, which may function as a protective factor. This is in line with previous studies that have shown that longer working hours and increasing job demands are associated with increases in burnout. [62,63]

Trait mindfulness scores in this sample spanned the full possible range of the FMI-14 [54], from 14-56, indicating variability in mindfulness levels among medical researchers. The mean score of 37.51 (SD=7.64), together with a median of 38 and a mode of 39, indicates that mindfulness levels in the sample were generally moderate and normally distributed. Taken together, these descriptive statistics indicate that most participants exhibited a moderate level of trait mindfulness.

Previous studies examining mindfulness in physicians, nurses, and other healthcare professionals have generally reported moderate mindfulness scores. [38–41] While physicians and researchers may differ in their day-to-day responsibilities, both groups operate in high-pressure environments marked by long hours, performance evaluation, and competing professional demands. The similarity in mindfulness distributions across these populations suggests that the protective role of mindfulness may generalize across clinical and non-clinical medical professions.

Moreover, the finding that younger participants reported lower mindfulness is in line with the literature suggesting age-related increases in mindfulness linked to improved emotional regulation and self-awareness. [64] Additionally, variations in mindfulness across regions may reflect distinct philosophical and religious foundations, with mindfulness remaining culturally grounded in Buddhist contemplative traditions in parts of Asia; while Western frameworks often emphasize mindfulness as a means of individual self-regulation and performance enhancement, influencing how the construct is understood and reported. [65]

Understanding burnout risk among medical researchers is critical, given their central role in advancing scientific knowledge and improving patient care indirectly through research outputs. Burnout in this population may have downstream consequences, including reduced research productivity, compromised research quality, and attrition from academic careers. [2,14,19,22,29–31,33] By situating medical researchers within the broader burnout literature, this study highlights that burnout is not confined to direct patient-care roles but is a systemic issue across medical and scientific disciplines, age groups, career stages, and geographic contexts. These findings also reinforce the multidimensional nature of burnout and highlight the importance of examining specific burnout domains to develop targeted interventions. Additionally, the moderate, negative correlation between burnout and mindfulness observed in the present sample is consistent with a growing body of literature demonstrating that higher mindfulness is associated with lower burnout and mindfulness-based interventions can potentially reduce burnout. [66–68] Extending this finding, hierarchical regression analyses indicated that mindfulness predicted burnout risk above and beyond sociodemographic factors. These results highlight mindfulness not only as a correlate but also as a potential protective factor against burnout, reinforcing the relevance of mindfulness-focused strategies in interventions aimed at promoting well-being and reducing burnout risk.

By examining burnout across diverse socio-demographic groups and in relation to trait mindfulness, this study provides a comprehensive assessment of burnout risk among medical researchers. Rather than isolating single factors, this approach highlights intersecting patterns of vulnerability. It offers a framework for future research to identify and investigate specific high-risk populations, such as demographic groups that have not yet been systematically studied, and to clarify the mechanisms underlying their elevated burnout risk. Future studies should also explore how individual-level factors, such as mindfulness and socio-demographic factors, interact with institutional and structural conditions to shape burnout risk among medical researchers.

### Strengths and Limitations

The study’s cross-sectional design ensured a relatively quick and cost-effective project, facilitated by online survey distribution, the absence of participant follow-up, and the lack of experimental manipulation. Additionally, the use of validated scales that measure trait burnout and mindfulness, rather than inferring these factors from qualitative data, allowed for a robust methodology and accurate analyses.

Additionally, in comparison to the Maslach Burnout Inventory (MBI) [69], the BAT-12 reflects a modern understanding of burnout as a work-related syndrome affecting emotional and cognitive functioning, uses clinically validated cut-off scores, and is open access for research purposes. [3,53] Similarly, employing statistical analysis to determine the relationship between variables further strengthened the credibility of our findings. Moreover, since the study was conducted on an international scale, these findings can be generalized to the broader medical research community. This is supported by our methodological framework that sampled diverse medical researchers whose work was published and indexed in MEDLINE and ensured a high response rate through reminder emails.

However, a limitation arose from the exclusion of non-English speaking researchers, as only journals published in English were sampled. Additionally, the survey was written and distributed in English. This limited the pool of potential participants and restricted the generalizability of findings to English-speaking medical researchers.

Furthermore, despite sending reminder emails and targeting recently published authors, some researchers may not have been able to respond due to external circumstances, such as retirement or short-term leave, potentially lowering the response rate. Moreover, the results only outline a possible correlation between burnout risk, trait mindfulness, and socio-demographic characteristics and cannot determine a causal relationship between them. The inherent limitations of a cross-sectional survey design, including susceptibility to recall bias and non-response bias, also remained. Furthermore, more comprehensive assessments of organizational factors may help clarify environmental influences on burnout beyond individual demographic characteristics.

## Conclusion

This study provided valuable insights into the prevalence of burnout risk among medical researchers and its association with trait mindfulness and socio-demographic predictors. While trait mindfulness scores varied, a substantial proportion of participants were at high or very high risk of burnout. Among these groups, mental distancing was the most prevalent, followed by exhaustion, emotional impairment, and cognitive impairment. A moderate to large, negative relationship was observed between burnout and mindfulness. Additionally, sex, age and employment status emerged as significant predictors for burnout, while age and region emerged as significant predictors for mindfulness. By leveraging an international survey and validated assessment tools, we generated evidence that can inform future interventions, coping strategies, and policies tailored to the unique challenges faced by medical researchers. While acknowledging the limitations of a cross-sectional design, these findings can contribute to a deeper understanding of burnout risk, trait mindfulness, and associated socio-demographic variables, potentially helping preserve the well-being and productivity of medical researchers globally.

## List of Abbreviations

BAT: Burnout Assessment Tool
BAT-12: 12-item version of the Burnout Assessment Tool
FMI: Freiburg Mindfulness Inventory
FMI-14: 14-item version of the Freiburg Mindfulness Inventory
REB: Research Ethics Board
OSF: Open Science Framework

## Declarations

### Ethics Approval and Consent to Participate

Ethics approval was sought and obtained from the University Hospital Tübingen Research Ethics Board (REB Number: 217/2025BO2) to conduct this study.

### Consent for Publication

All authors consented to this manuscript’s publication.

## Availability of Data and Materials

All relevant materials and data were included in this manuscript or posted on the Open Science Framework: https://doi.org/10.17605/OSF.IO/EAWD8

## Authors’ Contributions

JYN: designed and conceptualized the study, collected and analysed data, drafted the manuscript, and gave final approval of the version to be published.

NS: collected and analysed data, drafted the manuscript, and gave final approval of the version to be published.

GM: collected and analysed data, drafted the manuscript, and gave final approval of the version to be published.

MIB: analysed data, made critical revisions to the manuscript, and gave final approval of the version to be published.

AKK: provided input for the design and methodology, assisted with the collection and analysis of data, made critical revisions to the manuscript, and gave final approval of the version to be published.

HG: designed and conceptualized the study, assisted with the collection and analysis of data, made critical revisions to the manuscript, and gave final approval of the version to be published.

## Supplementary Files

**Appendix 1**: OVID MEDLINE Search Strategy Derived from All Journals Currently Indexed in MEDLINE

**Appendix 2**: Survey

**Appendix 3**: R Code Used in the Analyses

## Data Availability

All relevant materials and data were included in this manuscript or posted on the Open Science Framework.

https://doi.org/10.17605/OSF.IO/EAWD8

## Appendices

### Appendix 1: OVID MEDLINE Search Strategy Derived from All Journals Currently Indexed in MEDLINE

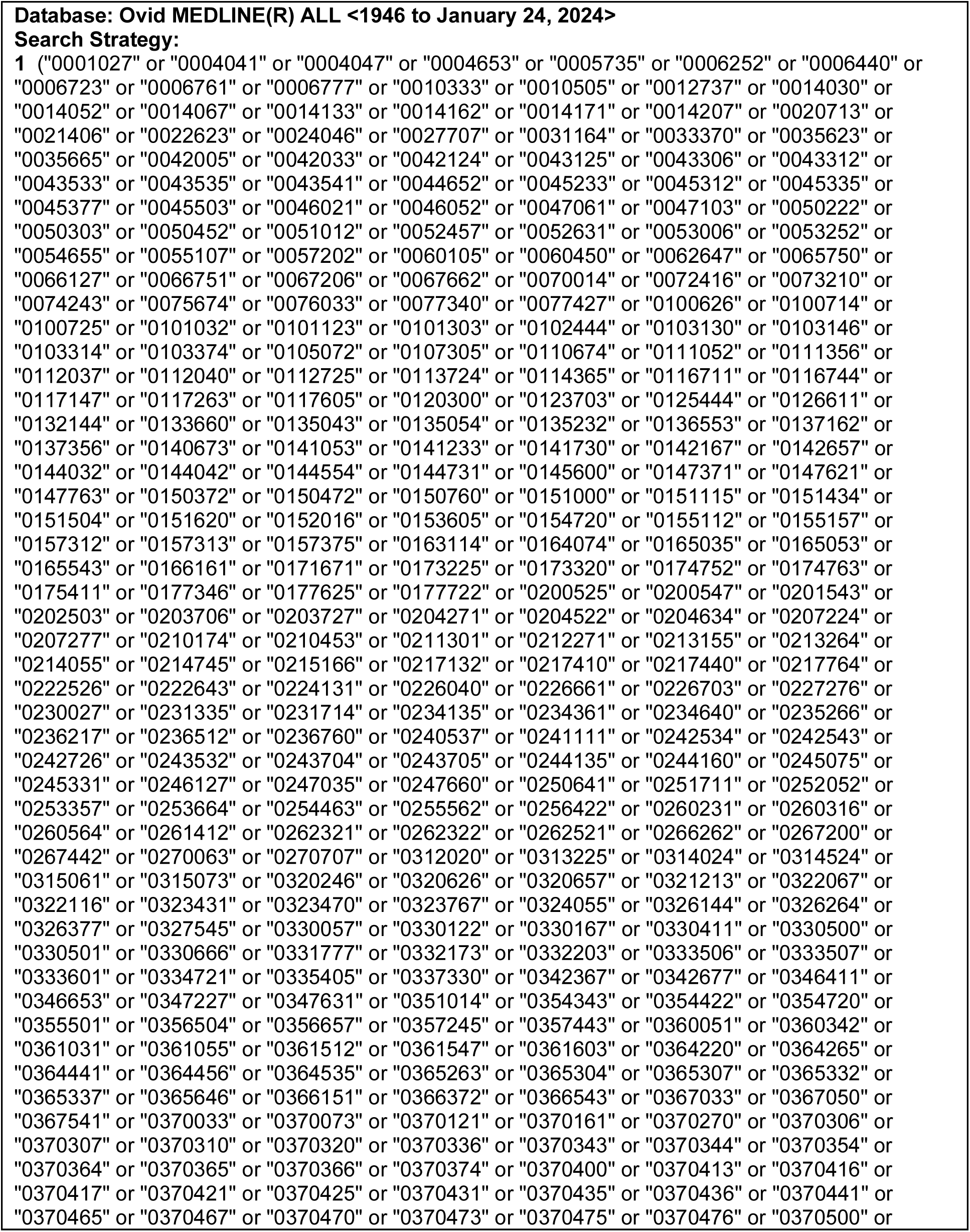
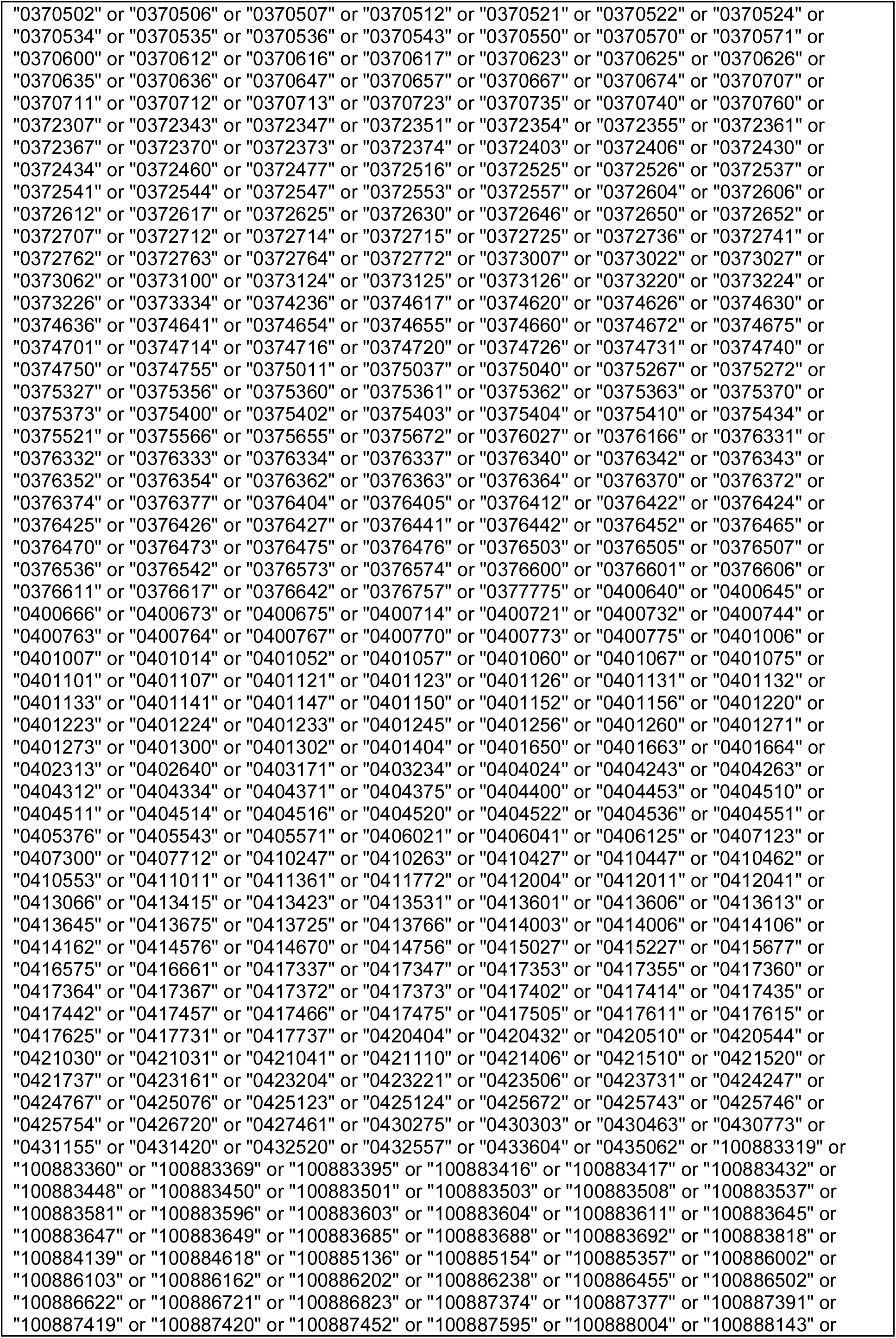
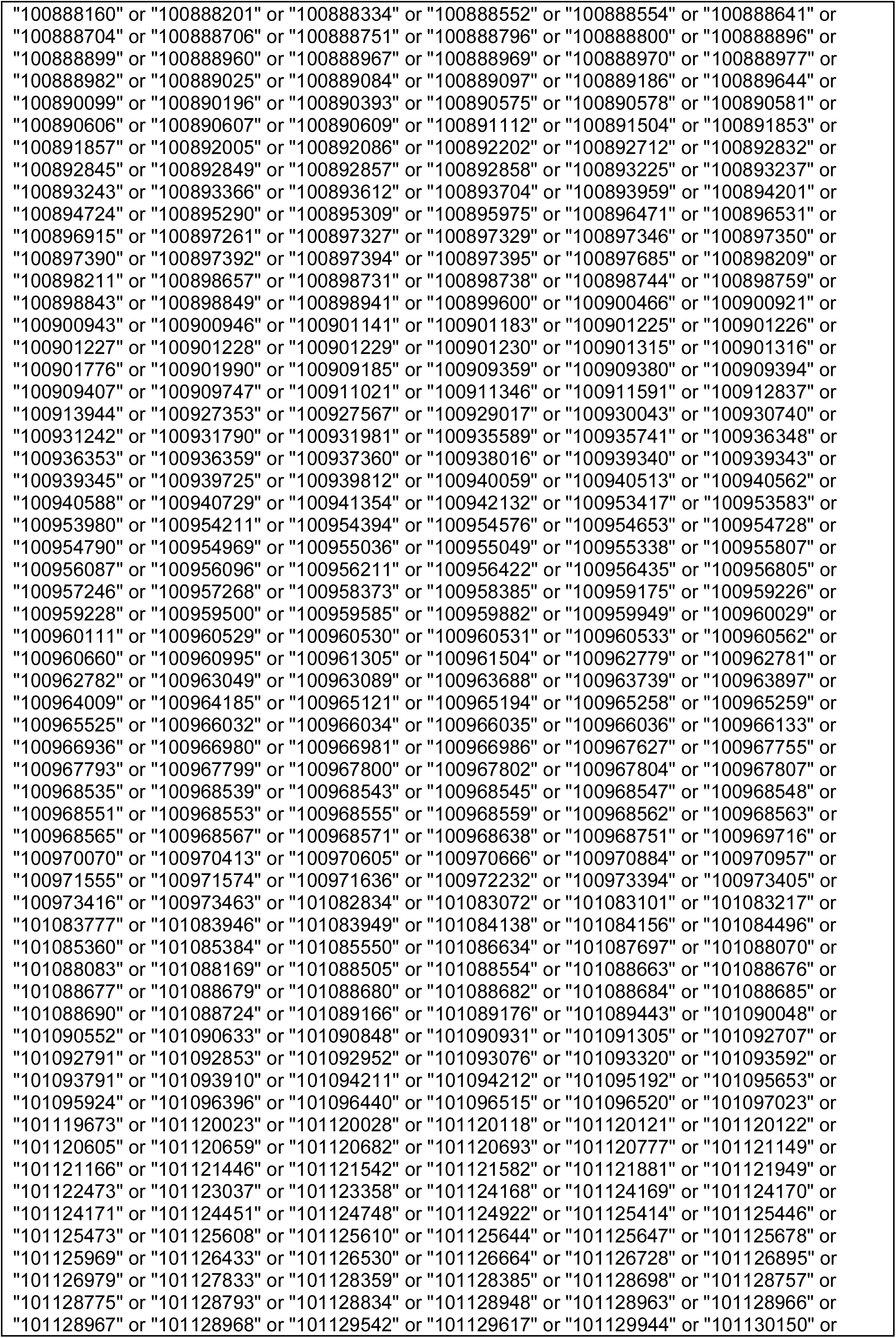
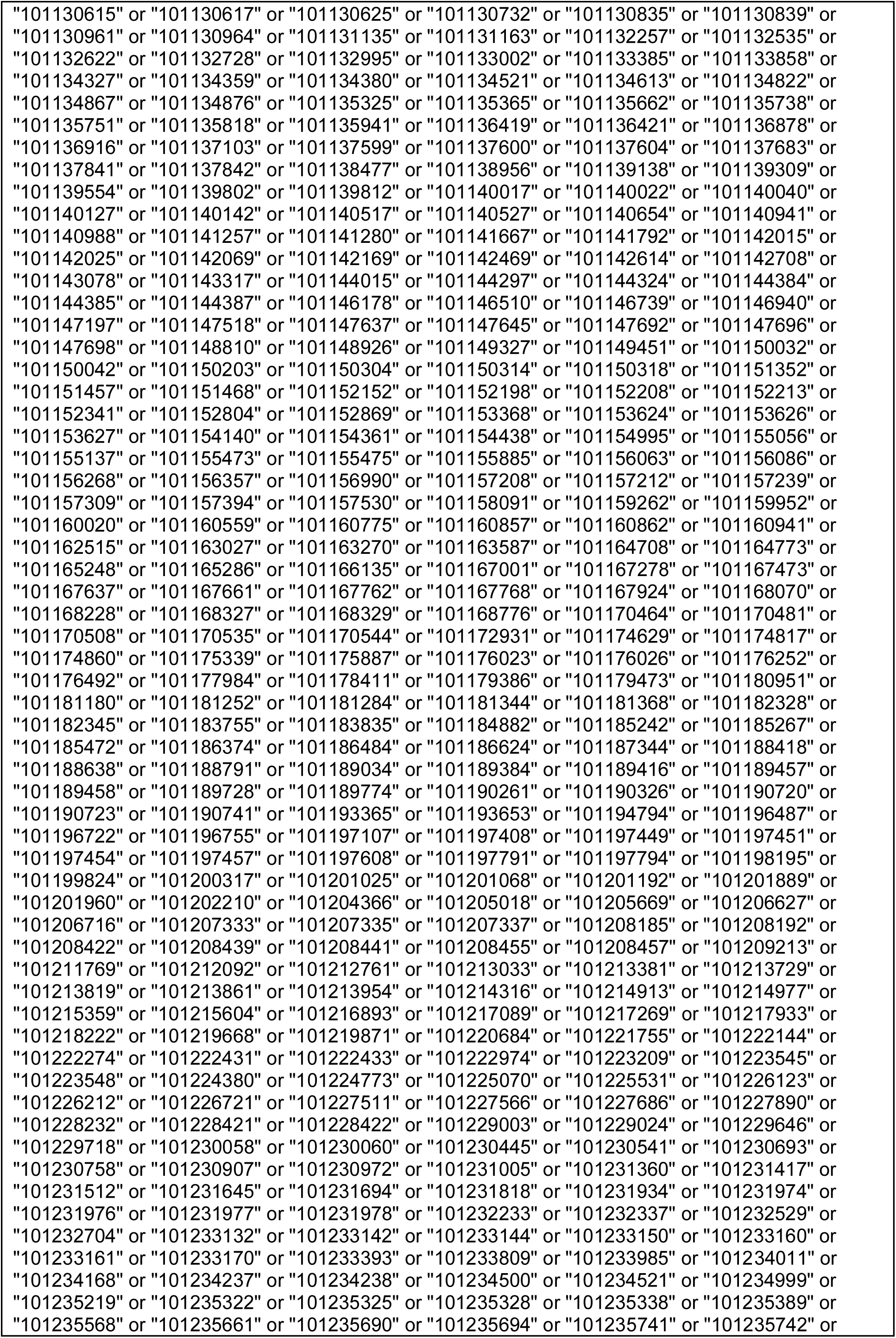
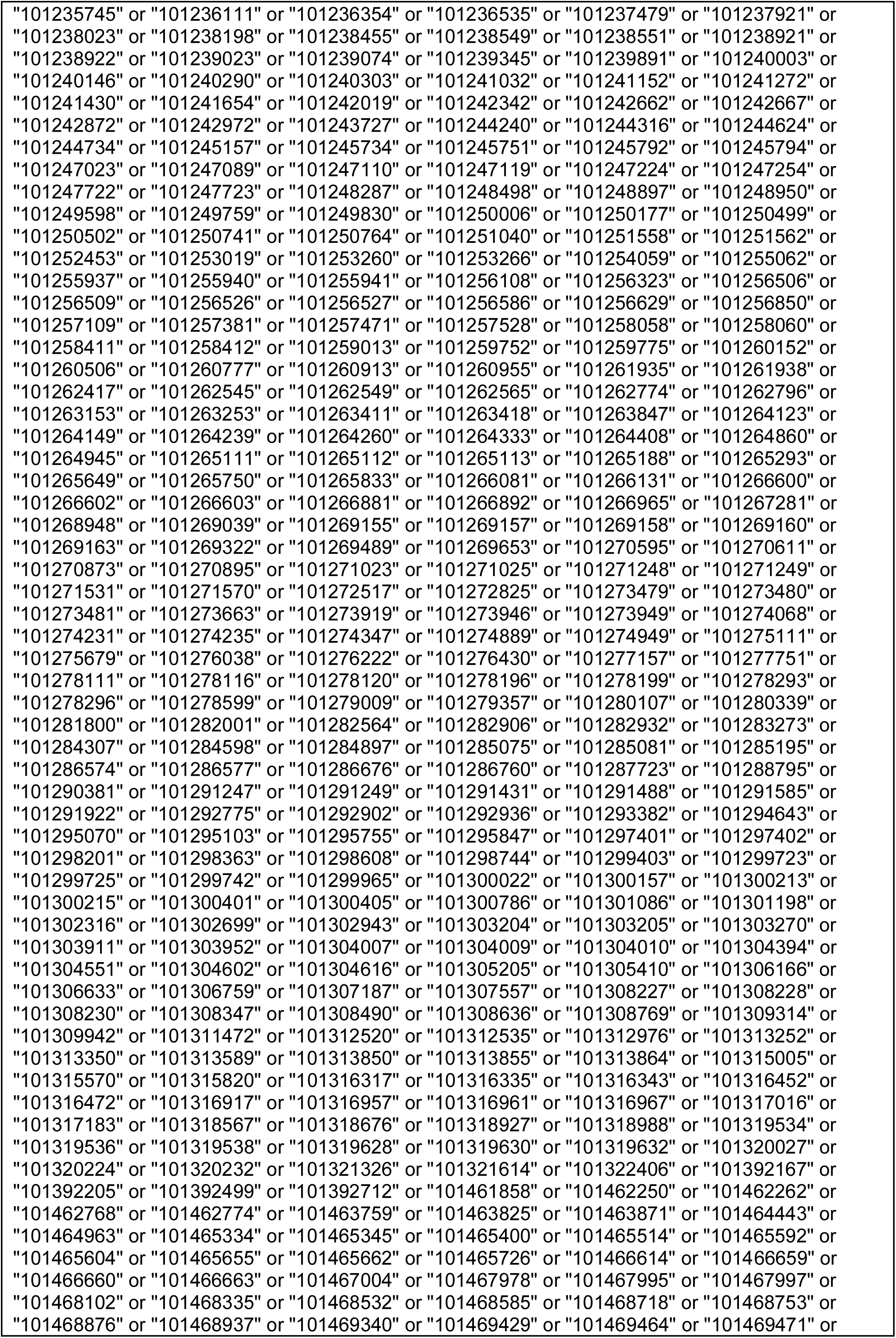
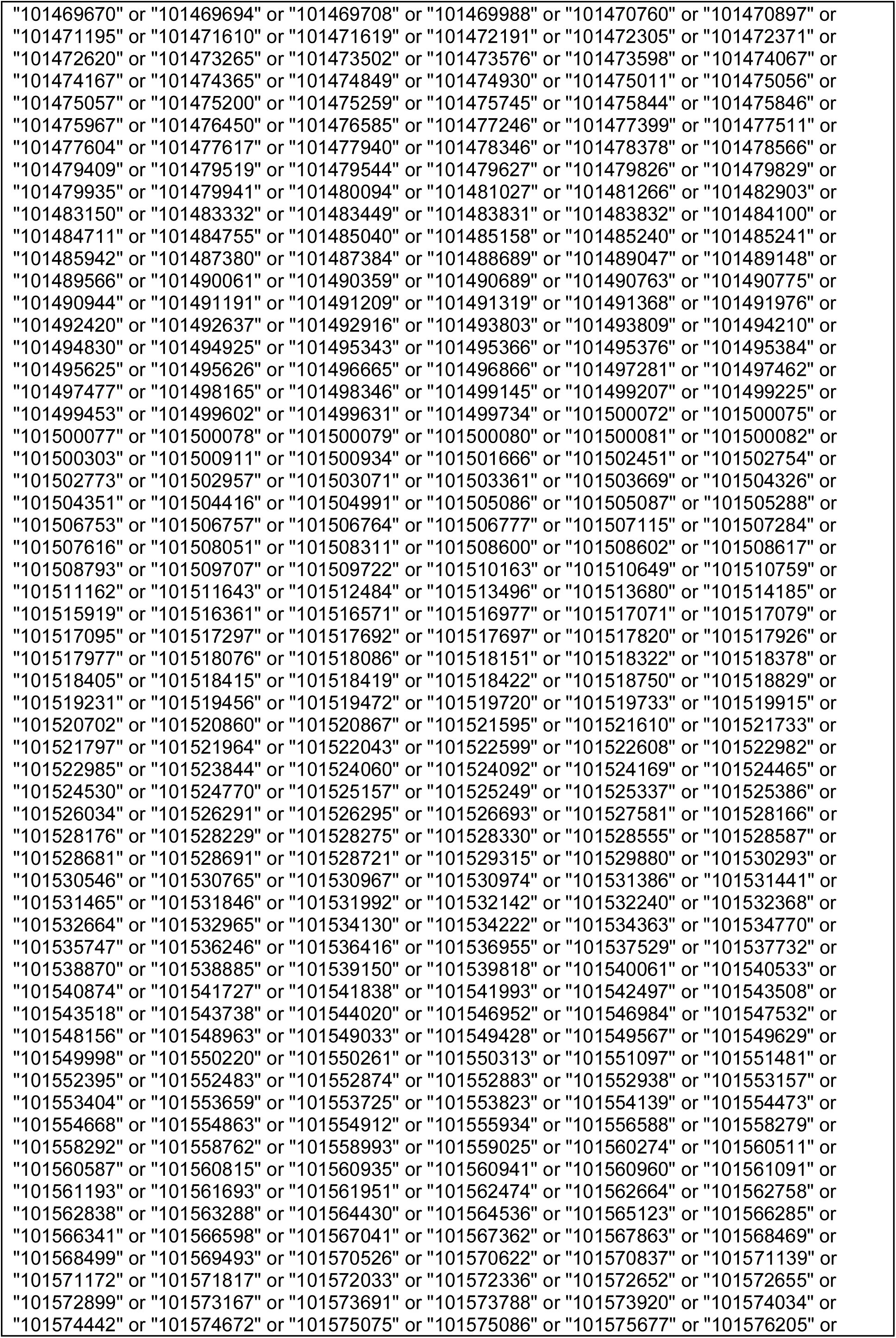
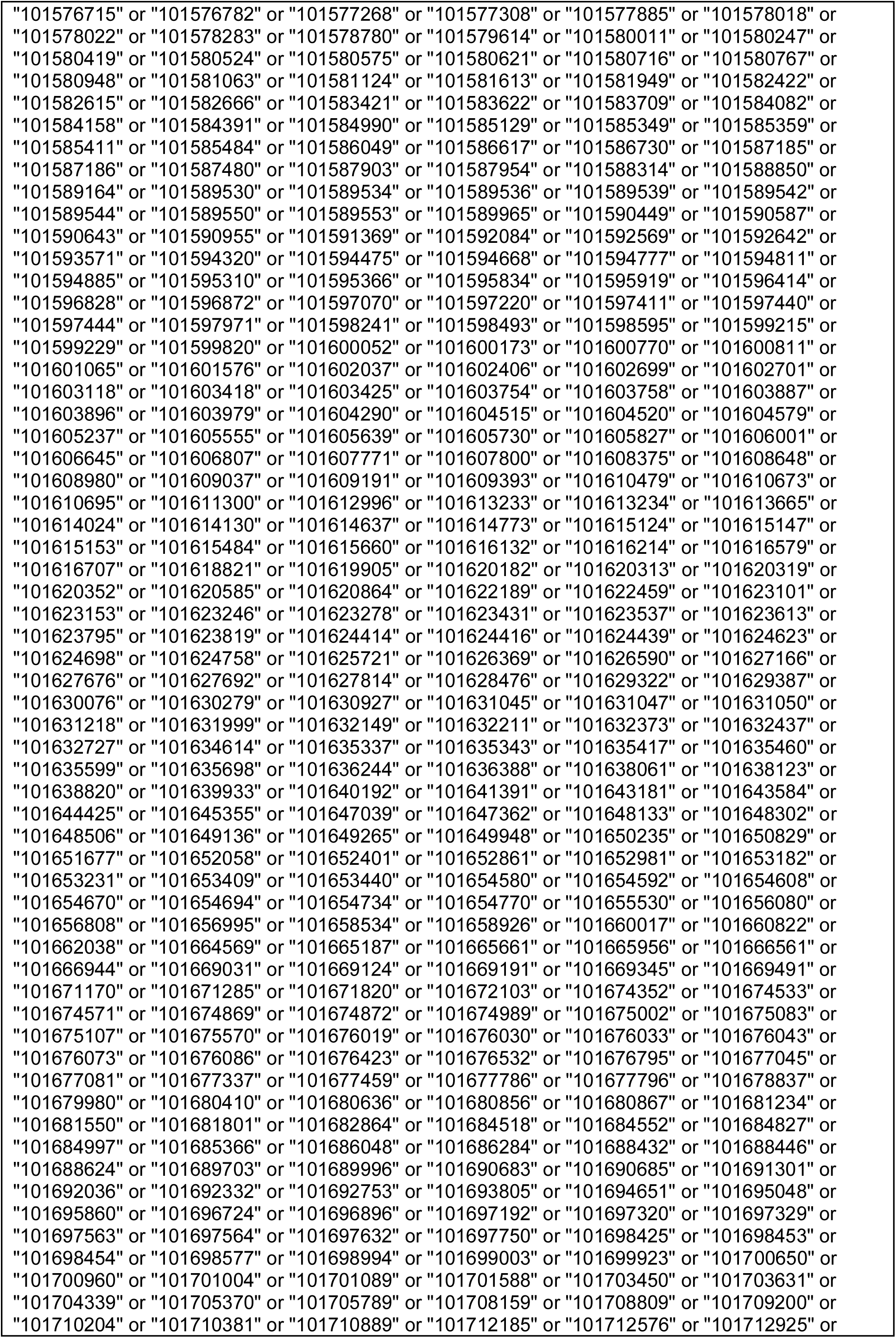
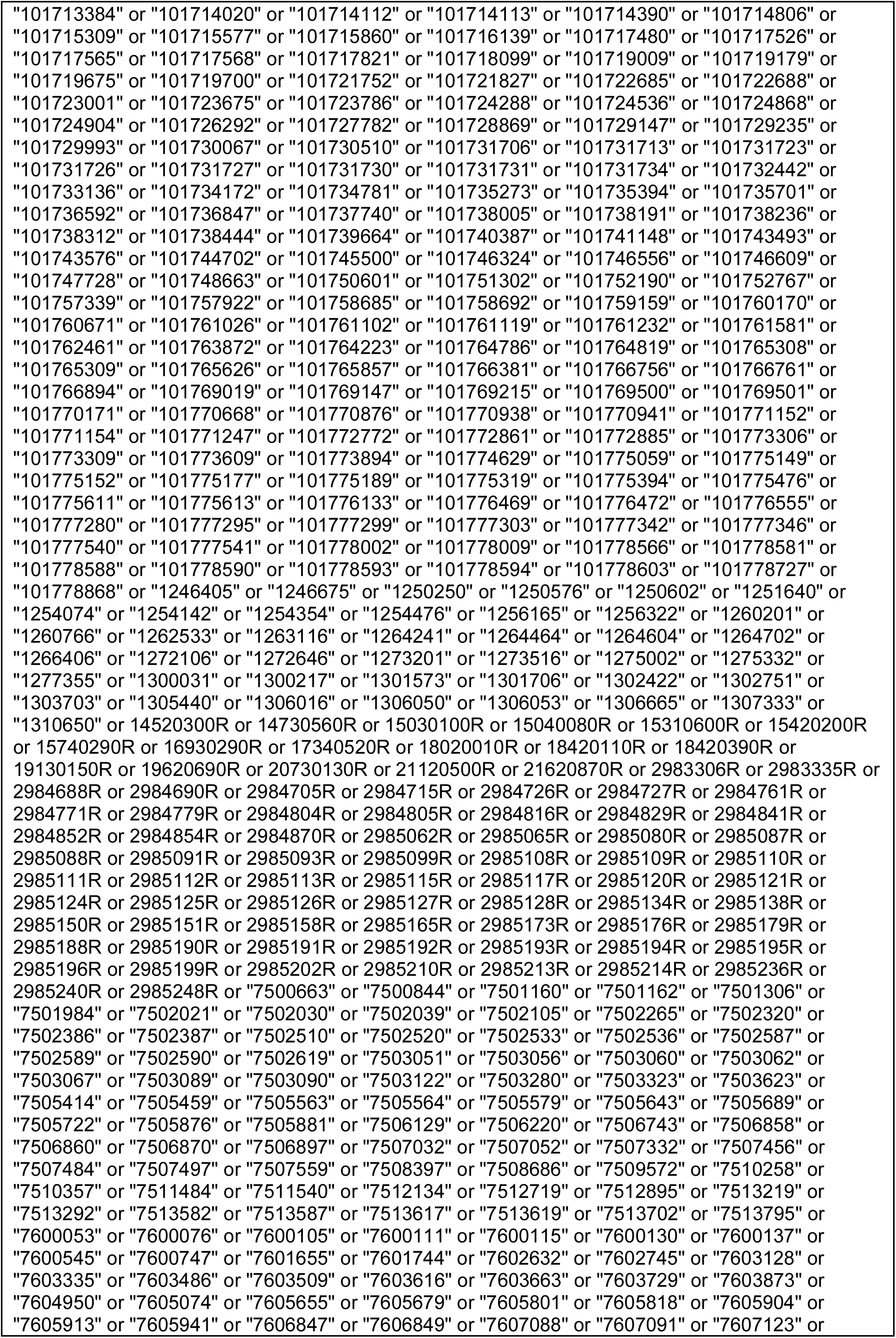
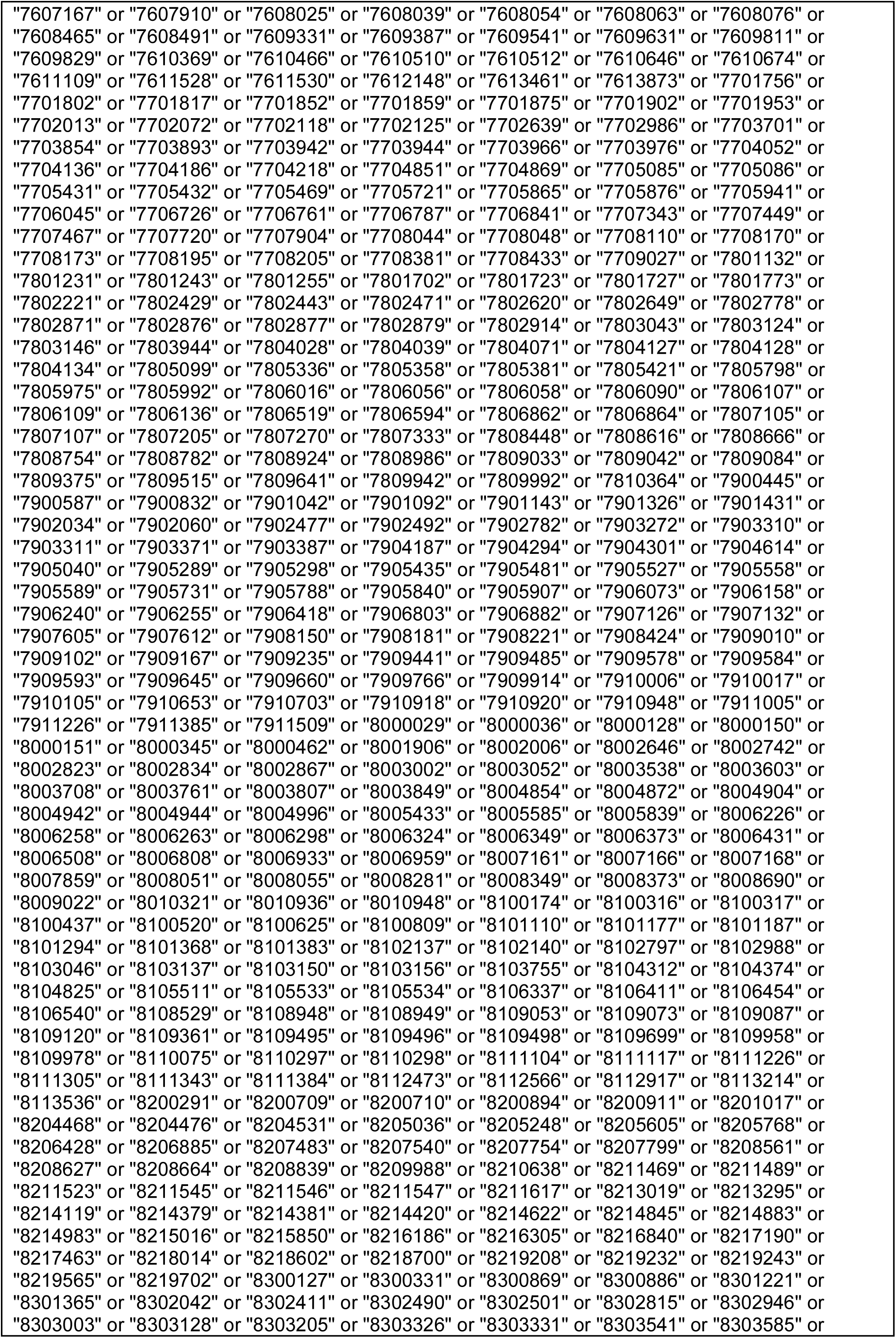
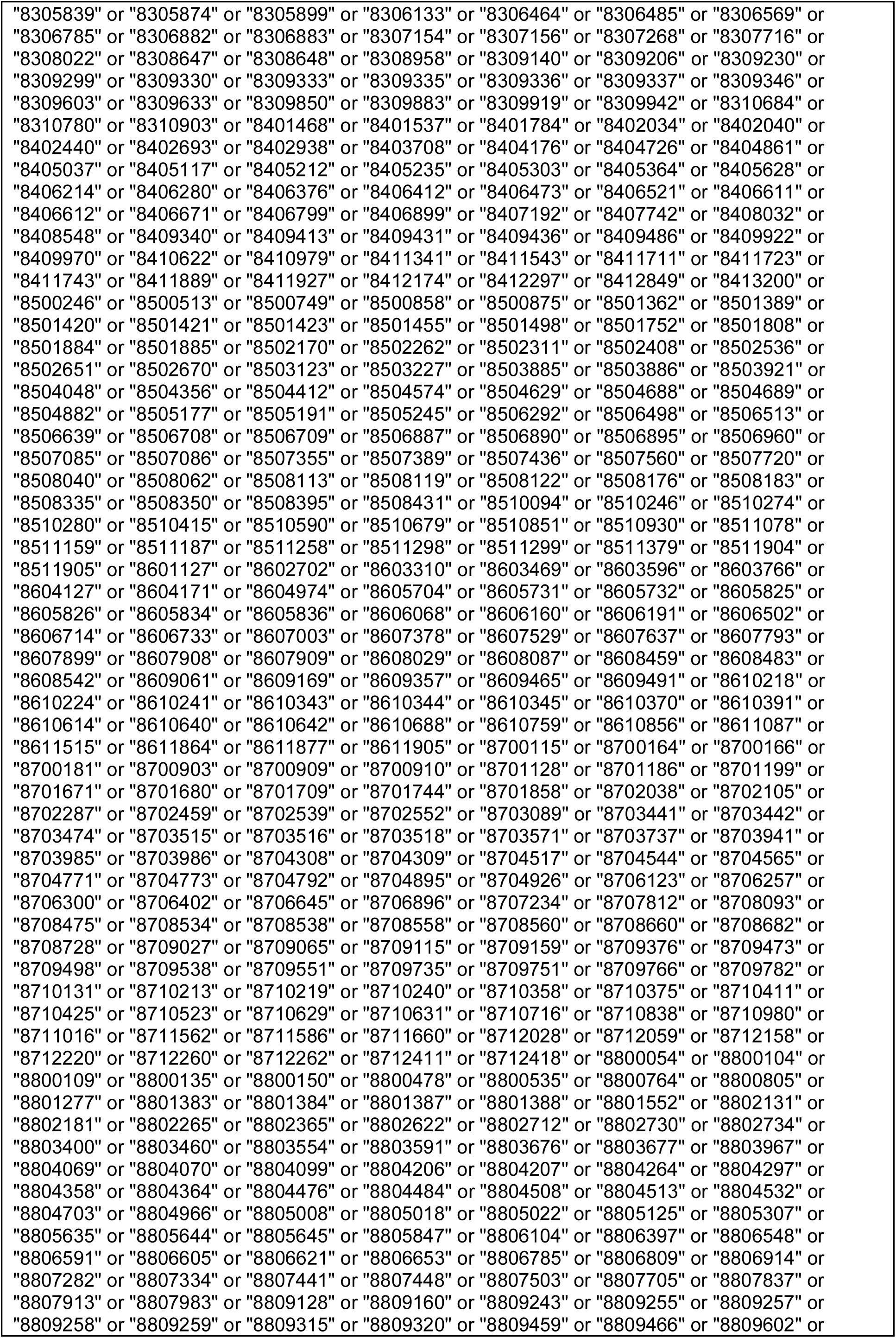
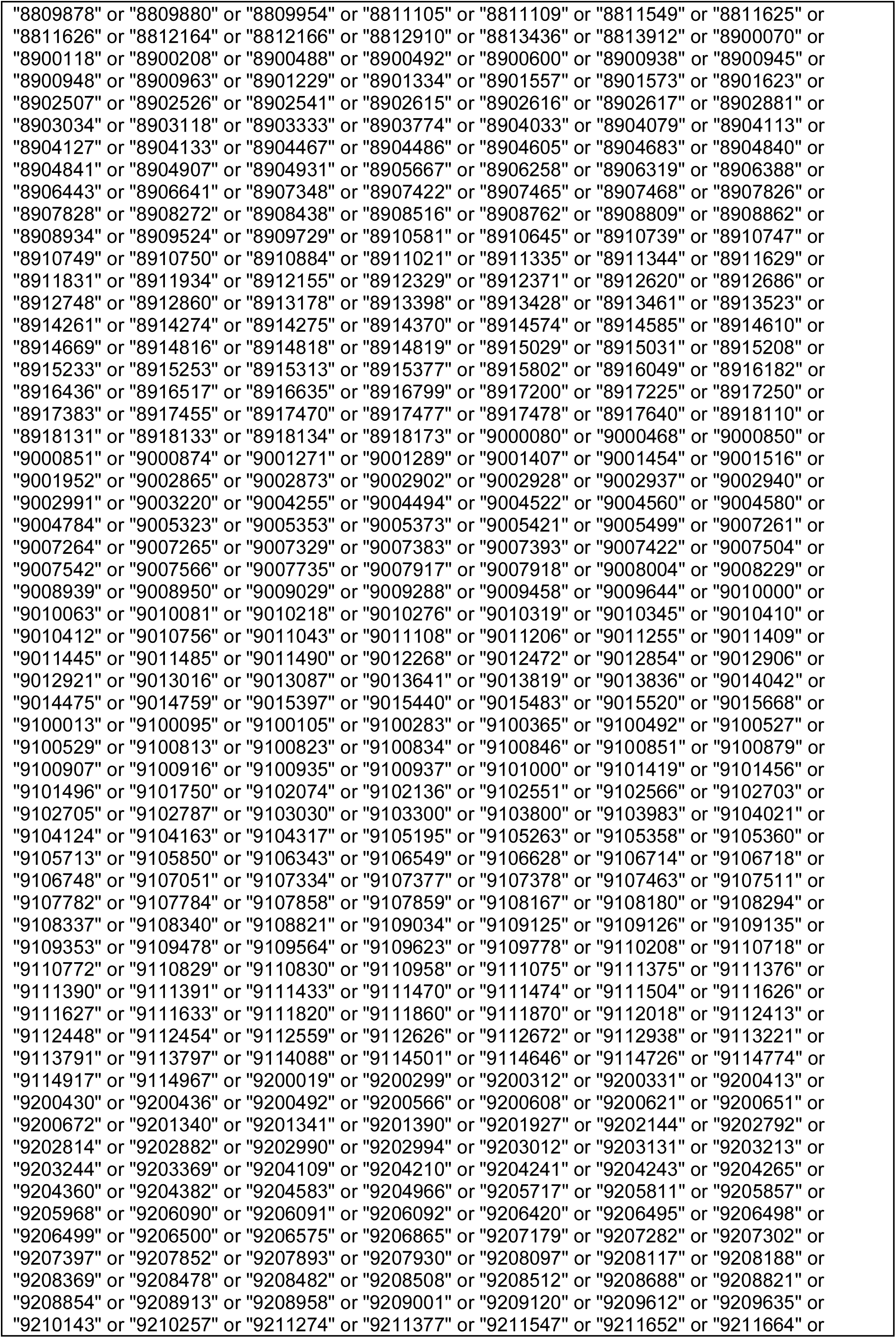
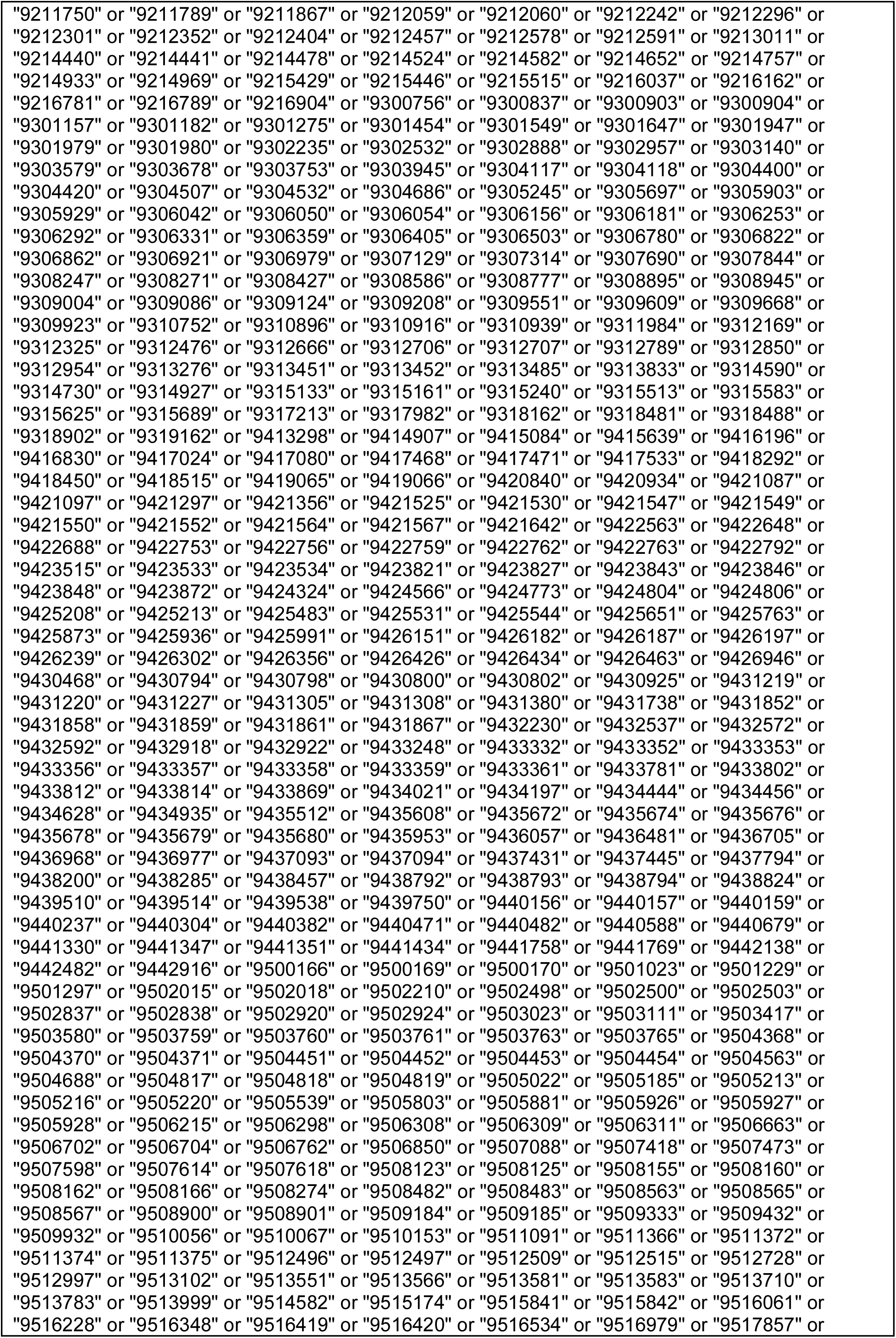
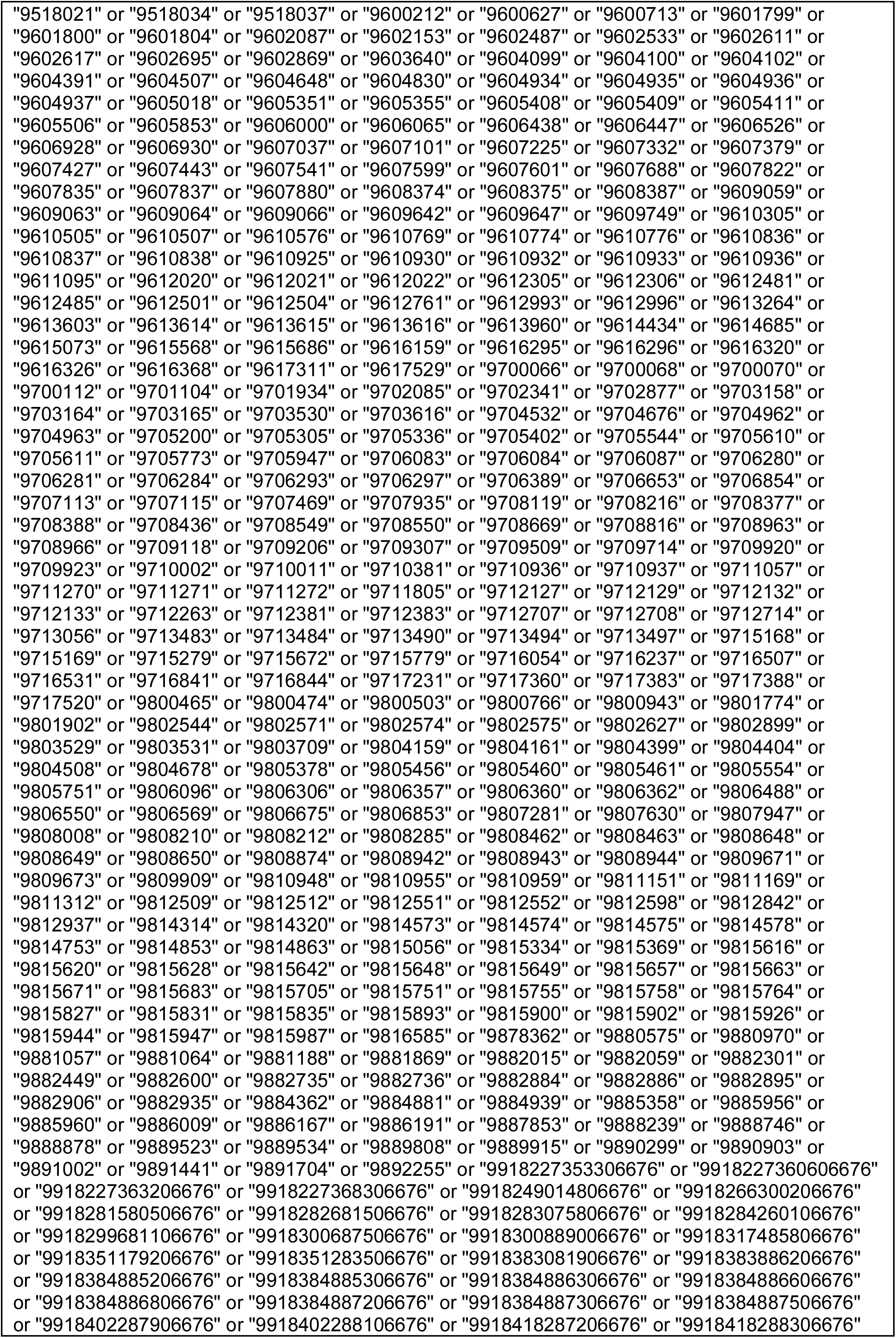
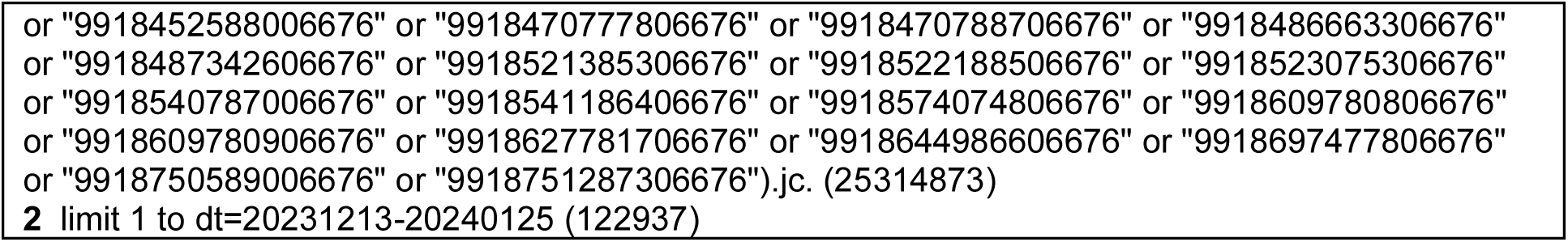

## Appendix 2: Survey

### Initial Screening Question

IMPORTANT NOTE: This survey is intended for participants who are medical researchers (of any kind, whereby your research in one way or another contributes to the field of medicine) who have completed at least one terminal degree in their respective field of study (e.g., PhD or equivalent in your respective field, MD or equivalent in your respective profession) OR have >5 years of experience in a research-focused role (e.g., research coordinator). If you do not meet these eligibility requirements (e.g., student with minimal research experience), you are ineligible to participate.

### Based on the above criteria, are you eligible to participate?

a. Yes
b. No

(Note: If the participant selects “No”, the survey ends.)

#### Demographic Questions

1. What career stage best describes you? (If you are currently pursuing a second degree following the completion of a terminal degree [e.g., PhD student after obtaining MD], please include the number of years since the completion of the terminal degree).

a. Early career researcher (<5 years of formally starting your career post PhD, MD, or an equivalent)

b. Mid-career researcher (5-10 years of starting your career post PhD, MD, or an equivalent)

c. Senior researcher (>10 years of starting your career post PhD, MD, or an equivalent)

d. Prefer not to disclose

2. What best describes your current position? (Please select all that apply.)

a. Faculty member at a university/academic institution

b. Academic research staff (e.g., research coordinator) at a university/academic institution)

c. Government researcher

d. Clinician researcher

e. Pharmaceutical industry/company researcher or employee

f. Scholarly communications employee (e.g., scholarly journals/publishing, medical writing)

g. Journal editor or editorial board member

h. Consultant (e.g., own or employed by research consulting firm)

i. Third sector (e.g., NGO, non-profit)

j. Prefer not to disclose

k. Other, please specify:

3. What is your sex?

a. Male

b. Female

c. Intersex

d. Prefer not to disclose

e. Prefer to self-describe, please specify.

4. What is your age?

a. <18

b. 18-24

c. 25-35

d. 36-45

e. 46-55

f. 56-65

g. 66-70

h. 71-75

i. 76-80

j. 80+

k. Prefer not to disclose

5. What World Health Organization World Region are you currently employed in? If you are employed in more than one region, please select the region of your primary employment. (See: https://www.who.int/countries)

a. Africa

b. Americas

c. Eastern Mediterranean

d. Europe

e. South-East Asia

f. Western Pacific

g. Prefer not to disclose

6. Is English your primary language?

a. Yes

b. No

c. Prefer not to disclose

7. Which of the following best describes your primary research area? (Please select all that apply.)

a. Clinical research

b. Preclinical research – in vivo

c. Preclinical research – in vitro

d. Health systems research

e. Health services research

f. Methods research

g. Epidemiological research

h. Prefer not to disclose

i. Other, please specify:

8. What best describes your research focus/discipline? Please select all that apply. (See: https://classification.nlm.nih.gov/outline)

a. Human Anatomy

b. Physiology

c. Biochemistry. Cell Biology and Genetics

d. Pharmacology

e. Microbiology and Immunology

f. Parasitology. Disease Vectors

g. Clinical Laboratory Pathology

h. Pathology

i. General Medicine. Health Professions

j. Public Health

k. Practice of Medicine

l. Communicable Diseases

m. Medicine in Selected Environments

n. Musculoskeletal System

o. Respiratory System

p. Cardiovascular System

q. Hemic and Lymphatic Systems

r. Digestive System

s. Urogenital System

t. Endocrine System

u. Nervous System

v. Psychiatry

w. Radiology. Diagnostic Imaging

x. Surgery. Wounds and Injuries

y. Gynecology. Andrology

z. Obstetrics

aa. Dermatology. Integumentary System bb. Pediatrics

cc. Geriatrics

dd. Dentistry. Oral Surgery ee. Otolaryngology

ff. Ophthalmology

gg. Hospitals and Other Health Facilities hh. Nursing

ii. History of Medicine. Medical Miscellany

9. What is your current employment status?

a. Full-time

b. Part-time

c. Self-employed

d. Unemployed

e. On leave

f. Prefer not to disclose

g. Other (please specify)

<u>Following question #9, participants will then be asked to complete the following validated questionnaires</u>:

1. **Burnout Assessment Tool (Work-Related Short Version, BAT-12)** Citation: Hadžibajramović E, Schaufeli W, De Witte H. Shortening of the Burnout Assessment Tool (BAT) from 23 to 12 items using content and Rasch analysis. BMC Public Health. 2022;22(1):560. doi:10.1186/s12889-022-12946-y
2. Friedberg Mindfulness Inventory (Short Version, FMI-14) Citation: Walach H, Buchheld N, Buttenmüller V, Kleinknecht N, Schmidt S. Measuring mindfulness—the Freiburg Mindfulness Inventory (FMI). Pers Individ Dif. 2006;40(8):1543–55. doi:10.1016/j.paid.2005.11.025

## Appendix 3: R Code Used in the Analyses

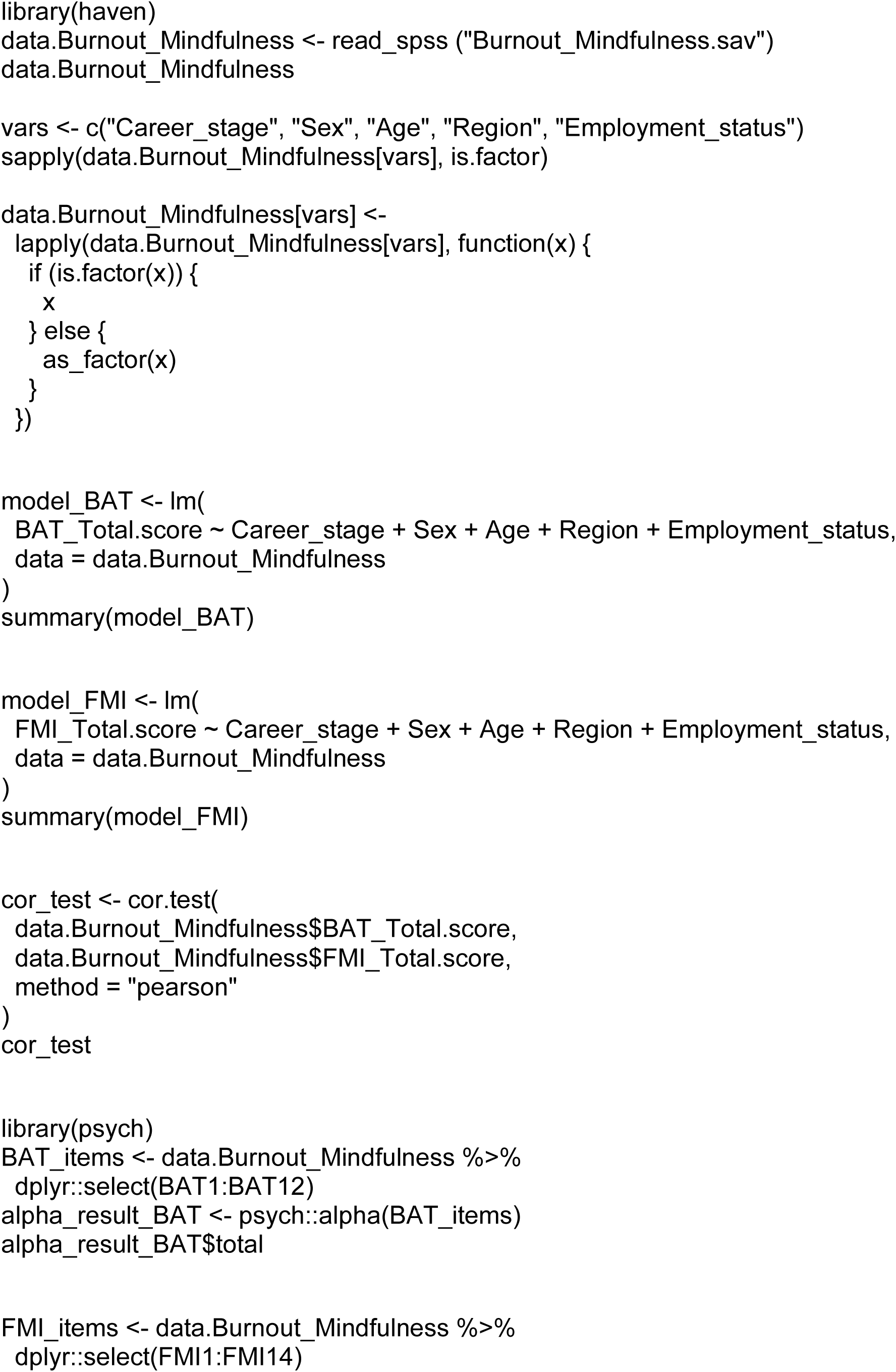
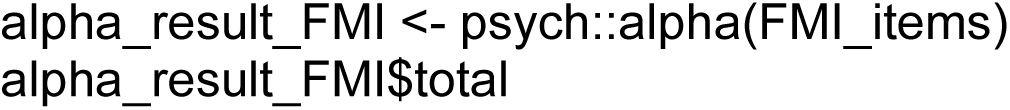

